# SARS-CoV-2 Detection in Istanbul Wastewater Treatment Plant Sludges

**DOI:** 10.1101/2020.05.12.20099358

**Authors:** Bilge Alpaslan Kocamemi, Halil Kurt, Ahmet Sait, Fahriye Sarac, Ahmet Mete Saatci, Bekir Pakdemirli

## Abstract

Following the announcement of SARS-CoV-2 worldwide pandemic spread by WHO on March 11, 2020, wastewater based epidemiology received great attention in several countries: The Netherlands [Medama et al., 2020; K-Lodder et al., 2020], USA [Wu et al., 2020; Memudryi et al., 2020], Australia [Ahmed et al., 2020], France [Wurtzer et al., 2020], China [Wang et al., 2020], Spain [Randazzo et al., 2020; Walter et al., 2020], Italy (La Rosa et al., 2020; Rimoldi et al., 2020) and Israel [Or et al., 2020], performed analysis in wastewaters by using different virus concentration techniques. Turkey took its place among these countries on 7^th^ of May, 2020 by reporting SARS-CoV-2 RT-qPCR levels at the inlet of seven (7) major municipal wastewater treatment plants (WWTPs) of Istanbul [Alpaslan Kocamemi et al., 2020], which is a metropole with 15.5 million inhabitants and a very high population density (2987 persons/km^2^) and having about 65 % of Covid-19 cases in Turkey. Sludges that are produced in WWTPs should be expected to contain SARS-CoV-2 virus as well. There has not yet been any study for the fate of SAR-CoV-2 in sludges generated from WWTPs. Knowledge about the existing of SARS-CoV-2 in sludge may be useful for handling the sludge during its dewatering, stabilizing and disposal processes. This information will also be valuable in case of sludges that are used as soil conditioners in agriculture or sent to landfill disposal.

In wastewater treatment plants, generally two different types of sludges are generated; primary sludge (PS) and waste activated sludge (WAS). PS forms during the settling of wastewater by gravity in the primary settling tanks. Little decomposition occurs during primary sludge formation. Since most of the inorganic part of the wastewater is removed in the earlier grit removal process, the PS consists of mainly organic material that settles. The PS is about 1-2 % solids by weight. In the biological treatment part of the WWTPs, the biomass that forms in the anaerobic, anoxic and oxic zones of the process is settled in final clarifiers by gravity and returned to the beginning of the biological process so that it is not washed off. The waste activated sludge (WAS) is the excess part of the biomass that grows in this secondary treatment process. It has to be removed from the process not to increase the mixed liquor suspended solids concentration (bacteria concentration) in the secondary process more than a fixed value. The WAS is about 0.6 - 0.9 % solids by weight.

This work aims to find whether SARS-CoV-19 is present in the PS and WAS before it is dewatered and sent to anaerobic or aerobic digester processes or to thermal drying operations.

For this purpose, on the 7th of May 2020, two (2) PS samples were collected from Ambarliı and Tuzla WWTPs, seven (7) WAS samples were collected from Terkos, Ambarliı, Atakoy I & II, Pasakoy II, Buyukcekmece and Tuzla I WWTPs. Polyethylene glycol 8000 (PEG 8000) adsorption [Wu et al., 2020] SARS-Cov-2 concentration method was used for SARS-CoV-2 concentration after optimization. [Alpaslan Kocamemi et al., 2020]. Real time RT-PCR diagnostic panel validated by US was used to quantify SARS-CoV-2 RNA in primary and waste activated sludge samples taken from WWTPs in Istanbul. All samples were tested positive. Titers of SARS-CoV-2 have been detected ranging copies between 1.17×10^4^ to 4.02×10^4^ per liter.

Value of the Data
The dataset provides information about SARS-CoV-19 in primary and waste activated sludges generated in WWTPs.
As being the first study in the world, the dataset presented is expected to be beneficial in handling the sludge during its dewatering, stabilizing and disposal processes

**Data Description:** SARS-CoV-2 copy numbers per liter measured for sludge samples from WWTPs were summarized in Table 1 and shown in Figure 1 together with SARS-CoV-2 copy numbers observed in an earlier study [Alpaslan Kocamemi et al., 2020] in the influent of the WWTPs from which the sludge samples were taken.

To the best of our knowledge, no study has yet been reported the presence of SARS-CoV-2 in primary sludge (PS) and waste activated sludge (WAS) samples. Herein we report the results of SARS-CoV-2 presence in two (2) PS and seven (7) WAS samples from WWTPs in Istanbul. A total of nine (9) sludge samples were collected on the 7th May of 2020 and investigated for presence of SARS-CoV-2 with RT-qPCR methodology. SARS-CoV-2 genome was detected quantitatively from all samples. Sludge samples presented CT ranging from 33.5 to 35.8. Titers of SARS-CoV-2 have been detected ranging from 1.17×10^4^ to 4.02×10^4^ per liter.

The detected numbers of SARS-CoV-2 in PS samples were found similar to those observed for WAS samples. SARS-CoV-2 copy numbers detected in PS and WAS on 7^th^ of May, 2020 are greater than the copy numbers observed in the influent of these WWTPs on 21^st^ April, 2020 [Alpaslan Kocamemi, 2020]. By considering the fact that the number of cases reported for Istanbul on the 7^th^ of May, 2020 is less than the cases reported for the 21^st^ April, 2020, it may be concluded that SARS-CoV-2 concentrations are more in both primary and waste activated sludge.

## Experimental Design, Materials and Methods

### 2.1 Sampling

The primary and waste activated sludge samples were collected from nine (9) WWTPs in Istanbul, Turkey. Figure 2 presents a map indicating the locations of these WWTPs. PS grab samples were obtained from Ambarliı and Tuzla-I WWTPs. Tuzla-I and Tuzla-II WWTPs have common primary sedimentation tanks. Although Atakoy I and Atakoy II WWTPs have separate primary sedimentation tanks, they were in maintenance and primary sludge from these two plants could not be sampled. Waste activated sludge (WAS) grab samples were brought from all the eight (8) WWTPs listed in Table 1. Paşaköy, Buyukcekmece and Terkos plants do not have primary sedimentation therefore, only WAS was sampled from these plants. Because a portion of the virus can be lost to the supernatant, both PS and WAS samples were grab samples taken before the sludges were dewatered, by gravity thickeners or by mechanical dewatering units. Although both sludge samples contain 98-99 % water, filtering the sludge samples caused frequent clogging of the 0.45 and 0.2 micron filters. Therefore, the protocol used in raw wastewater samples was modified for the sludge samples.

**Figure 1.**
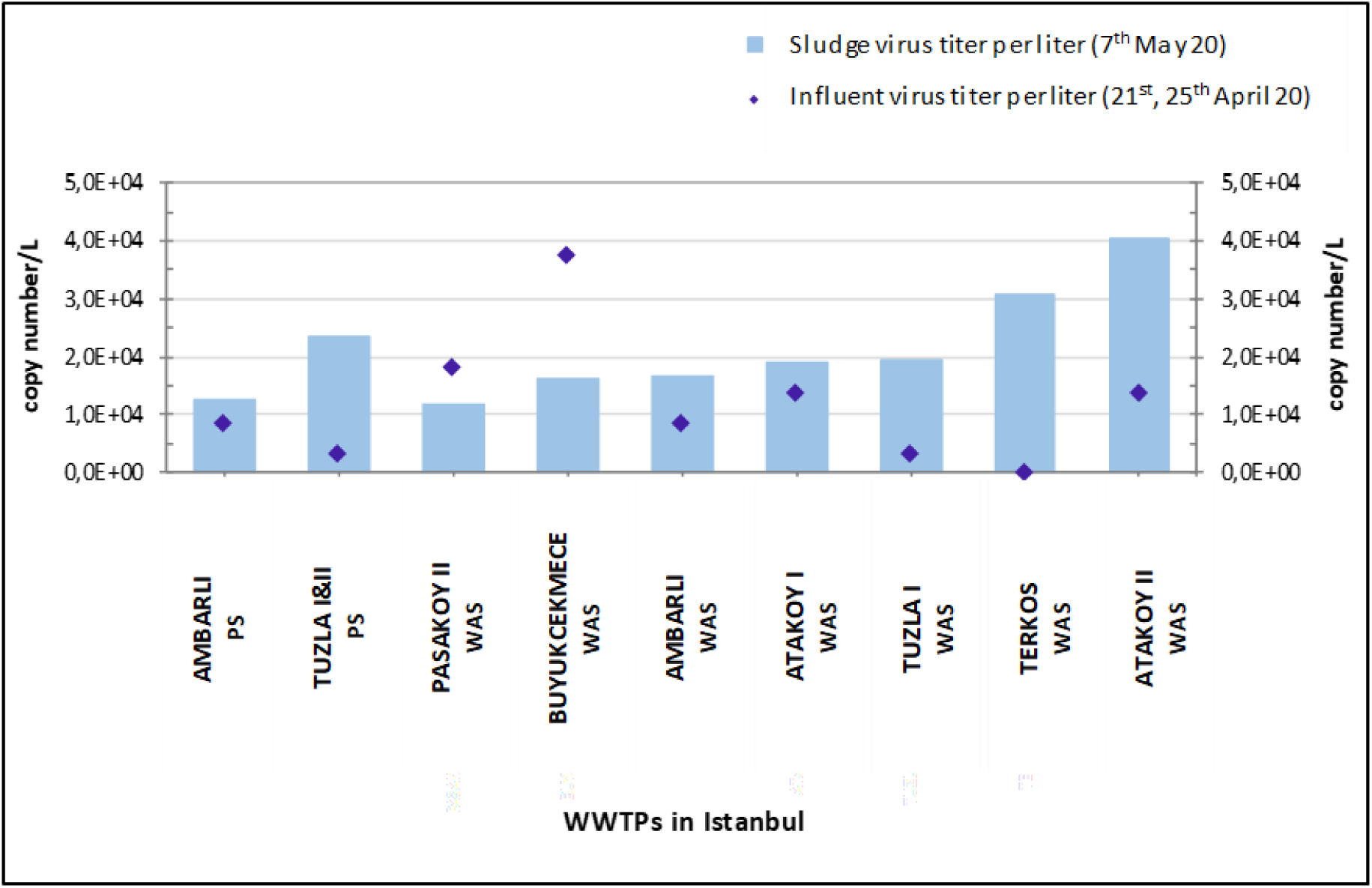
SAR-CoV-2 Levels in primary and waste activated sludges of Istanbul WWTPs.

**Figure 2.**
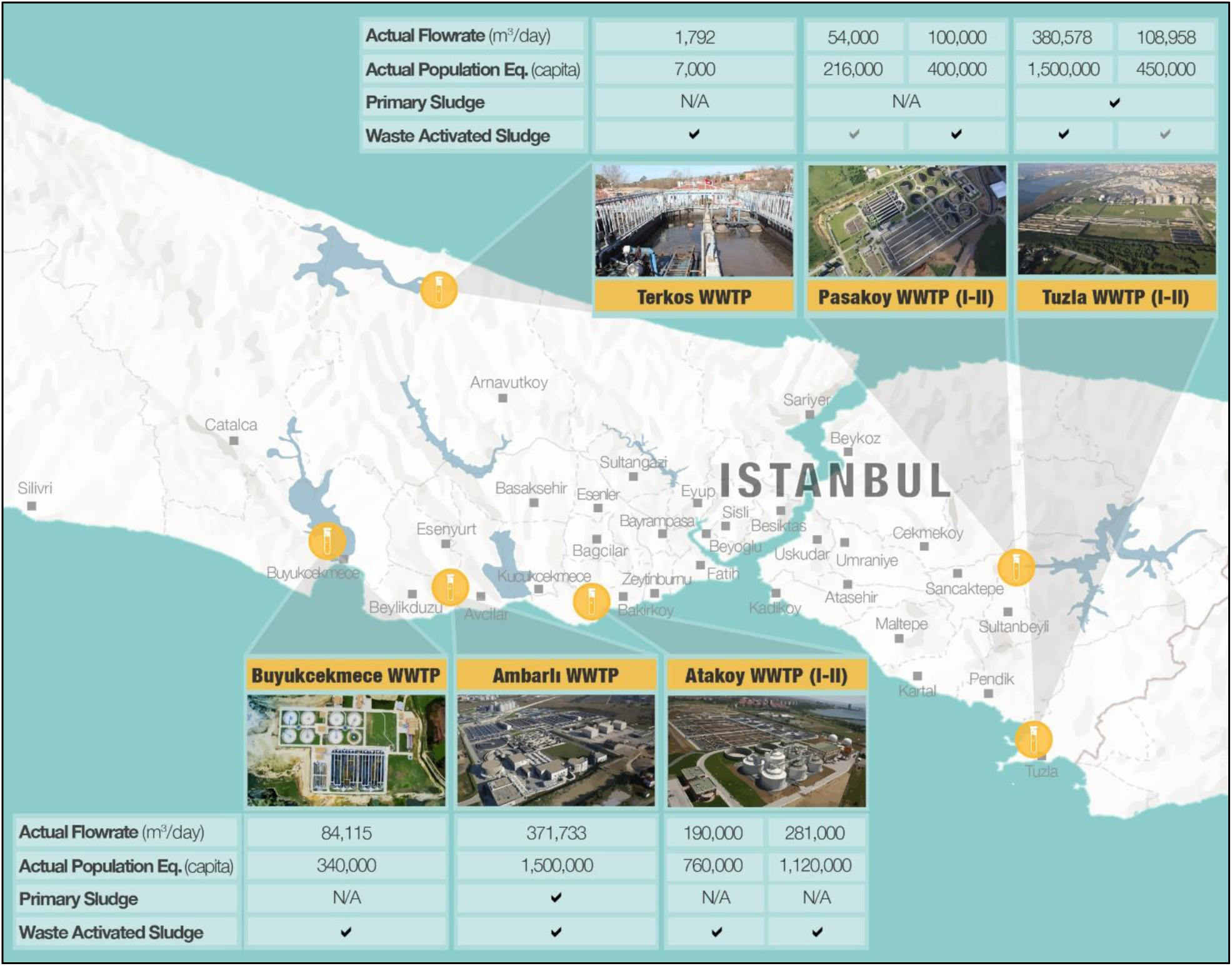
Primary and waste sludge sampling WWTPs in Istanbul

**Table 1.** SARS-CoV-2 RT-qPCR results of sludges taken from Istanbul WWTPs

| Sampling Point | Cq value | Cq SD | RT-qPCR results | RT-qPCR SD | Sludge virus titer/L | Virus titer per liter SD | slope | Reaction efficiency % | R<sup>2</sup> |
| --- | --- | --- | --- | --- | --- | --- | --- | --- | --- |
| **AMBARLI WWTP**<br>*Primary Sludge (PS)* | 35.86 | 1.60513 | 1.57E+03 | 1.41E+03 | 1.25E+04 | 1.12E+04 | -3.7 | 93.2 | 0.99 |
| **TUZLA I&II**<br>*Primary Sludge* | 34.71 | 0.51695 | 2.91E+03 | 8.60E+02 | 2.33E+04 | 6.88E+03 | -3.7 | 93.2 | 0.99 |
| **PASAKOY II WWTP**<br>*Waste Activated Sludge (WAS)* | 35.67 | 0.82024 | 1.47E+03 | 7.54E+02 | 1.17E+04 | 6.03E+03 | -3.7 | 93.2 | 0.99 |
| **BUYUKCEKMECE**<br>*Waste Activated Sludge (WAS)* | 35.00 | 0.27185 | 2.03E+03 | 3.52E+02 | 1.62E+04 | 2.82E+03 | -3.7 | 93.2 | 0.99 |
| **AMBARLI**<br>*Waste Activated Sludge (WAS)* | 34.98 | 0.26907 | 2.05E+03 | 3.34E+02 | 1.64E+04 | 2.67E+03 | -3.7 | 93.2 | 0.99 |
| **ATAKOY I**<br>*Waste Activated Sludge (WAS)* | 34.735 | 0.27577 | 2.39E+03 | 4.10E+02 | 1.91E+04 | 1.13E+04 | -3.7 | 93.2 | 0.99 |
| **TUZLA I WWTP**<br>*Waste Activated Sludge (WAS)* | 34.61 | 0.28378 | 2.44E+03 | 2.00E+02 | 1.95E+04 | 1.60E+03 | -3.7 | 93.2 | 0.99 |
| **TERKOS WWTP**<br>*Waste Activated Sludge (WAS)* | 34.11 | 0.13435 | 3.86E+03 | 3.46E+02 | 3.08E+04 | 2.77E+03 | -3.7 | 93.2 | 0.99 |
| **ATAKOY II**<br>*Waste Activated Sludge (WAS)* | 33.52 | 0.02828 | 5.03E+03 | 9.19E+01 | 4.02E+04 | 7.35E+02 | -3.7 | 93.2 | 0.99 |

The flowrates received by the WWTPs during 7^th^ of May 2020 are listed in Table 2. During the time of sampling (7^th^ of May, 2020), most of the industrial and commercial sites were shut down due to restrictions and the wastewater received by the plants were mostly domestic wastewater. The equivalent population served by the treatment plants was estimated using the 250 liters per day per capita value advised by İstanbul Water and Sewerage Administration (iSKi). The hydraulic retention time (HRT) and the sludge retention time (SRT) values of the WWTPs during the day the sludges were sampled are presented Table 3. HRT values were added to the table since they show the approximate washout time of the influent. The SRT values indicate whether the sludge is stable during the time of sampling.

**Table 2.**
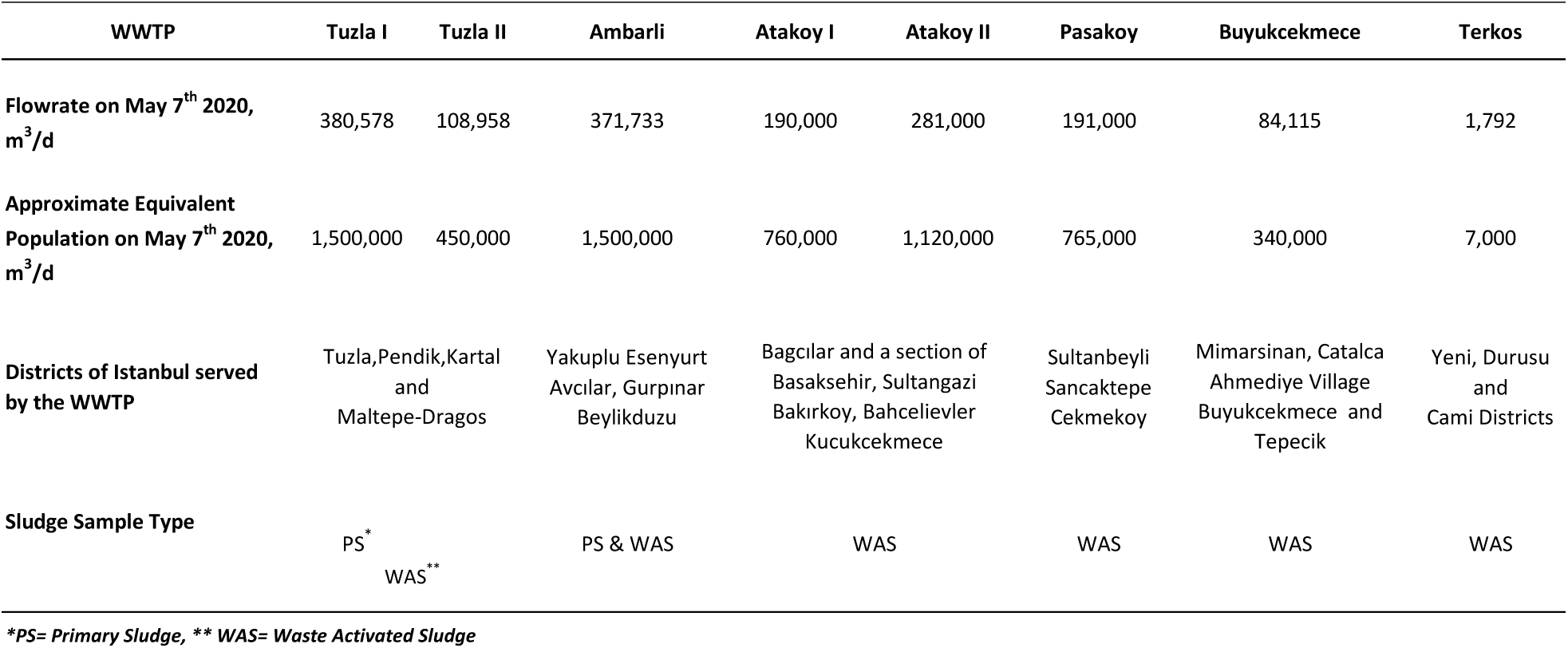
WWTP flowrates during the time of sludge sampling

**Table 3.** HRT and SRT values of the WWTPs during the day of sludge sampling

| WWTP | Tuzla I | Tuzla II | Ambarli | Atakoy I | Ataköy II | Pasakoy II | Buyukcekmece | Terkos |
| --- | --- | --- | --- | --- | --- | --- | --- | --- |
| Flowrate on May 7<sup>th</sup> 2020, m<sup>3</sup>/d | 380,578 | 108,958 | 371,733 | 190,000 | 281,000 | 191,000 | 84,115 | 1,792 |
| Wastewater Temperature, °C on May 7<sup>th</sup> 2020 | 19 | 19 | 19.5 | 21.5 | 21.1 | 20 | 19 | 18 |
| SRT<sub>stable</sub>, days (ATV-131, Eqn 5-5) | 15.4 | 15.4 | 14.8 | 12.9 | 13.3 | 14.3 | 15.4 | 16.5 |
| SRT on May 7<sup>th</sup> 2020, days | 8.3 | 18 | 16 | 26 | 12 | 12 | 15 | 20 |
| HRT on May 7<sup>th</sup> 2020, hours | 1.5 | 11.9 | 17 | 33 | 16.1 | 10.9 | 21.1 | 4 |

## Tuzla-I and Tuzla-II Wastewater Treatment Plants

Tuzla-I WWTP is designed for carbon removal and nitrification. It does not have biological phosphorous removal and denitrification units. Tuzla tertiary WWTP (Tuzla-II) is designed for carbon, nitrogen and biological phosphorous removal. Tuzla-I and Tuzla-II plants have common primary sedimentation tanks. Both plants treat the wastewater collected from Tuzla, Pendik, Kartal and Maltepe (Dragos) districts of istanbul. The treated wastewater is discharged into the Marmara Sea.

## Ambarlı Wastewater Treatment Plant

Ambarlı tertiary WWTP treats wastewater collected from Arnavutköy, Avcılar, Başakşehir, Beylikduzu, and Esenyurt districts of Istanbul covering and area of 438 km^2^. The treated wastewater is discharged into Haramidere Creek which discharges into the Marmara Sea.

## Atakoy-I and Atakoy-II Wastewater Treatment Plants

Atakoy I and II tertiary WWTPs treat wastewater from Bağcılar, Bahçelievler, Bakırköy, Başakşehir, Esenler, Küçükçekmece and Sultangazi districts of İstanbul. The treated wastewater is used for irrigation of recreational areas and in the industry. The effluent goes into Ayamama Creek which discharges into the Marmara Sea.

## Paşakoy-II Wastewater Treatment Plant

Pasakoy-II tertiary WWTP treats wastewater from Sultanbeyli, Sancaktepe, Çekmeköy districts of İstanbul. Pasakoy-II WWTP does not have primary sedimentation and anaerobic digesters. The waste activated sludge is stabilized using extended aeration process. Carbon, nitrogen and phosphorous are removed through anaerobic, anoxic and oxic stages (A^2^/O process). After deep sand filtration and UV disinfection, the treated wastewater is discharged into the Riva Creek which joins the Black Sea.

### 2.2 SARS-CoV-2 concentration

Prior to viral concentration steps, samples were shaken at 4°C at 100 rpm for 30 min to transfer viruses to the aqueous phase. Bacterial debris and large particles were removed from the samples by centrifugation at 7471G for 30 minutes at 4°C (Hettich, Rotanta 460R, Tuttlingen, Germany). Then, 250 ml of supernatant was filtered through 0.45 μm and 0.2 μm to remove remaining particles. A 5X PEG 8000 Stock Solution was prepared by dissolving 100 g of PEG 8000 (Nordic Chemical Solutions, Norway) and 17.5 g of NaCl in 200 ml distilled water. The pH was adjusted to 7.0-7.2 and the solution was sterilized with 0.2 μm filter. Filtrate was mixed thoroughly with PEG 8000 (10% w/v) by shaking for 1 minute. The mixture was incubated at 4oC at 100 rpm for overnight. Following to the incubation, the mixture was divided in six 50 ml falcon tubes. Viruses were precipitated by centrifugation at 7471G for 120 minutes at 4oC (Hettich, Rotanta 460R, Tuttlingen, Germany). Supernatant was removed carefully without disturbing the pellets. Pellets of each falcon tubes were re-suspended with 200 μl RNA free water. 1 ml of virus concentrate was used for total RNA extraction and remaining concentrate stored at −80°C. Total viral RNA was extracted with Roche MagNA pure LC total nucleic acid isolation kit using Roche MagNA pure LC system (Penzberg, Germany) in accordance with the manufacturer’s protocols. RNA were determined both qualitatively and quantitatively by Thermo NanoDrop 2000c (Penzberg, Germany).

### 2.3. Quantitative reverse transcription PCR (RT-qPCR)

Primers and taqman probe sets targeting SARS-CoV-2 RdRp gene were used in this study (Corman et al. 2020) to detect and quantify SARS-CoV-2 virus. Serial dilution of synthetic SARS-CoV-2 RdRp gene were used as a standard for absolute quantification. RT-qPCR analysis was performed in Realtime ready RNA virus Master (Roche Diaonostics, Mannhaim, Germany) contained 0.8 nM of forward primer and reverse primer, 0.25 nM probe and 5 μL of template RNA. The RT-qPCR assays were performed at 50 °C for 6 min, 53 °C for 4 min, 58 °C for 4 min for reverse transcription, followed by 95 °C for 1 min and then 45 cycles of 95 °C for 10 s, 58 °C for 30 s and 72 °C for 1 s for data collection using a Roche LightCycle 2.0 thermal cycler (Roche Diaonostics, Mannhaim, Germany).

## Data Availability

The authors confirm that the data supporting the findings of this study are available within the article.

## Ethics Statement

The work did not involve any human subject and animal experiments.

## Acknowledgments

This work was financed by Republic of Turkey, Ministry of Agriculture and Forestry.

The authors wish to acknowledge the Turkish Water Institute (SUEN) for the coordination and execution of this study. We wish to express our appreciation to the Ministry’s Veterinary Control and Research Institute for their hard work and rapid analysis of the wastewater samples. We thank to DSI (State Hydraulic Works) for their logistic support for intercity sample transportation. We also thank to ISKI (Istanbul Water and Sewerage Administration) for their cooperation and efforts to collect and preserve the sewage samples rapidly. Our special thanks to Dr. Esra Erdim from Marmara University Department of Environmental Engineering for her contribution to gather and collate up to date information about the worldwide studies on SARS-CoV-2 in wastewater.

